# Transformer-based structuring of Italian electronic health records with application in cardiac settings

**DOI:** 10.64898/2026.01.22.26344603

**Authors:** Sara Mazzucato, Andrea Bandini, Daniele Sartiano, Giuseppe Vergaro, Stefano Dalmiani, Michele Emdin, Silvestro Micera, Calogero Maria Oddo, Claudio Passino, Sara Moccia

## Abstract

**Purpose:** Natural Language Processing (NLP) has the potential to extract structured clinical knowledge from unstructured Electronic Health Records (EHRs). However, the limited availability of annotated datasets for algorithm training restricts its application in clinical practice. This study investigates the use of transformer-based NLP models to structure Italian EHRs in cardiac settings, addressing this gap.

**Methods:** We implemented and evaluated three named entity recognition algorithms: SpaCy, Flair, and Multiconer. The experiments utilized three datasets comprising 2235 anamneses from patients at the Fondazione Toscana Gabriele Monasterio, Italy.

**Results:** The SpaCy model achieved the highest performance with an F1-score of 97% in identifying clinical features on explicitly mentioned entities (Presence/Absence classification). However, features are not always mentioned, as clinicians selectively document only clinically relevant information in real-world practice. External validation shows model generalizability: EVD-100 dataset (considering 12 features, 97.13% F1) and STEMI dataset (considering 3 shared features, 88.29% F1). These structured variables were subsequently used to train machine learning algorithms (Logistic Regression, XGBoost, CatBoost) for classifying amyloidosis in heart failure patients. The classifiers trained on SpaCy-structured data attained an average F1-score of 66.70%, closely matching the 66.99% F1-score from classifiers using clinician-annotated data.

**Conclusion:** This study shows the feasibility of using NLP for structuring Italian EHRs in realistic clinical settings, highlighting its potential to enhance computer-assisted detection despite selective documentation patterns. The comparable performance across annotation methods suggests NLP’s capability to bridge the gap in dataset annotation, paving the way for its integration into clinical practice.

## 1 Introduction

In the last decades, the availability of Electronic Health Records (EHRs) in medicine has increased significantly, providing a comprehensive perspective on patients’ health statuses and enabling clinicians to anticipate the detection of a wide range of diseases [1]. However, unstructured EHRs present challenges due to their volume and complexity, requiring time-consuming manual extraction [2]. Efficient access to structured clinical information is crucial for timely diagnosis, especially for key conditions like hypertension and cardiac diseases [3].

Machine Learning (ML) can enhance early detection but is limited by unstructured clinical narratives. Natural Language Processing (NLP) can convert these texts into structured data to aid clinical decisions. While NLP methods are advanced in English [4], Italian clinical NLP remains underdeveloped due to scarce datasets [5].

This study proposes a pipeline using transformer-based Named Entity Recognition (NER) models, SpaCy, Flair, and Multiconer, to structure Italian EHRs. Transformers capture lexical variability better than rule-based methods [6], and SpaCy is noted for strong sequence labeling [**?**]. Flair and BERT have proven effective in English medical NLP [7].

The dataset includes 405 anonymized anamneses from Fondazione Toscana Gabriele Monasterio (FTGM), with 200 patients having Cardiac Amyloidosis (AM), a rare and challenging diagnosis [8, 9]. SpaCy was also validated on two external datasets totaling 1830 records from various cardiac conditions.

Our goals are to develop a robust NLP pipeline for Italian EHRs and evaluate NER models for AM patient classification using the extracted features. Key contributions are:

1. Applying transformer-based NER models to Italian cardiology EHRs for the first time.
2. Validating these models on three real-world datasets totaling 2235 patients.

This work aims to advance Italian clinical NLP and support automated patient care through structured data extraction.

## 2 State of the art

In the rapidly evolving field of EHR structuring, various NLP methodologies have been proposed for cardiac applications, highlighting the growing interest in clinical NLP [1, 10].

The study of Choi et al. [11] introduces a Graph Convolutional Transformer (GCT) for heart failure identification on 91,026 English EHRs (AUCPR = 0.86), though GCT may underperform on short and variable-length texts [12]. Jin et al. [13] compare LSTM and CNN models for heart failure risk prediction, showing LSTM superiority, although they can struggle with sparse signals in long anamneses [14].

Garvin et al. [15] extract the Ejection Fraction from echocardiogram reports using rule-based NLP, achieving high performance (F1 = 0.99), but focusing only on a single feature. Similarly, Nagamine et al. [16] apply word embeddings to extract concepts from Russian EHRs (AUC = 0.68), though such methods may falter with long clinical texts [12].

The study of Annetta et al. [17] adopts BERT for processing over 50,000 Polish EHRs, achieving 0.70 accuracy. Likewise, Silvert et al. [18] identify amyloidosis signs via BERT-based models, achieving performance close to clinician labels. These works inspire our study of transformer models for Italian EHRs, offering advantages in handling long and sparse narratives [19]. The diversity of NLP approaches in EHR structuring for cardiac settings reflects the ongoing exploration of innovative solutions. Here, we specifically focus on the innovative application of NER in structuring Italian anamneses.

## 3 Methods

In this section, we present the datasets and the methods used for EHR structuring. Our pipeline is shown in Fig. 1. The detailed flowchart of the workflow is in Fig. 2.

**Fig. 1.**
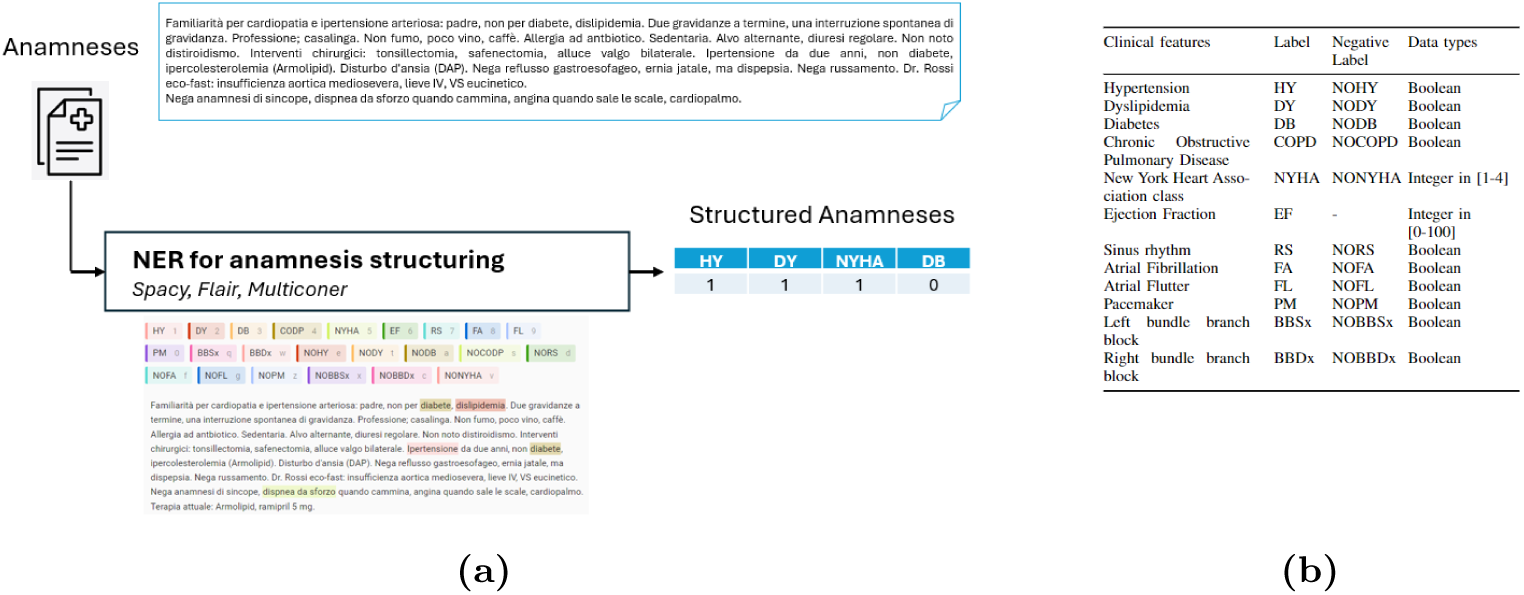
(a) Proposed pipeline: the textual anamneses are processed using Named Entity Recognition (NER) models (SpaCy, Flair, Multiconer) to identify clinical features. An example of a manually annotated anamnesis using Label Studio annotation software is reported and different colors correspond to different labels (b) Clinical features labels and data type

### 3.1 Datasets

We used three databases from FTGM: the AM dataset for training, and EVD-100 and STEMI datasets for external validation.

The AM dataset includes anonymized anamneses from 405 patients (2010-2023): 200 with amyloidosis (AM, 75% male, age 77 ± 9 years) and 205 without (noAM, 63% male, age 73 ± 11 years). It contains 11,098 sentences and 124,742 tokens, with an average anamnesis length of 248 words. We intentionally used a balanced dataset to ensure sufficient examples for machine learning classification. Each anamnesis represents a clinical summary documenting the patient’s medical history and current status as recorded by physicians at the time of evaluation. The extracted features reflect the clinical information documented at that specific timepoint and do not capture temporal evolution of comorbidities across multiple visits.

Twelve clinical features were manually annotated using LabelStudio^1^, creating a gold standard dataset based on relevant literature [20], including both positive and negative labels.

For external validation, the EVD-100 dataset (100 anamneses, FTGM Cardiothoracic Department) provided annotations for all 12 features. The STEMI dataset (1,730 patients, 24% female, age 75 ± 11 years) was annotated only for three shared features: hypertension, dyslipidemia, and diabetes.

Table 1 provides comprehensive statistics. The distribution of anamnesis lengths is shown in Appendix in Figure 7.

The study was approved by FTGM Ethics Committee (Decree No. 3854, 02/12/2023) and conducted per Declaration of Helsinki. All participants provided informed consent. Clinical records were anonymized at source by FTGM staff prior to data sharing, following GDPR-compliant protocols.

**Fig. 2.**
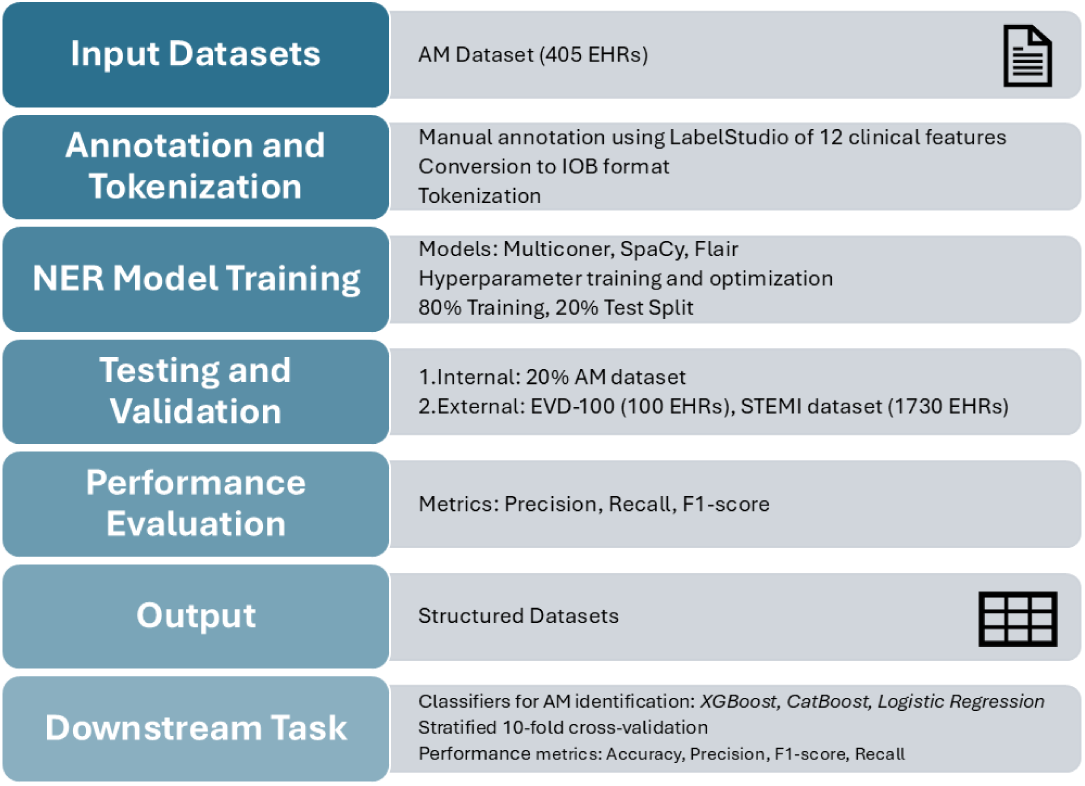
Flowchart of the workflow for structuring Electronic Health Records (EHRs) The process begins with the collection of input datasets, followed by the annotation and tokenization of clinical information Then, Named Entity Recognition (NER) models are trained, and their results are tested and validated Performance evaluation includes key metrics such as precision, recall, and F1-score Finally, a downstream task is conducted using different classification algorithms

**Table 1.** Descriptive statistics for the three clinical databases used in this study.

| Characteristic | AM dataset | EVD-100 | STEMI |
| --- | --- | --- | --- |
| Number of records | 405 | 100 | 1,730 |
| Avg. words per anamnesis | 256 (12–1,113) | 188 (7–383) | 202 (2–348) |
| Avg. characters per anamnesis | 1,869 (70–8,206) | 1,357 (47–2,517) | 1,455 (29–2,047) |
| Total sentences | 11,469 | 1,520 | 28,956 |
| Total tokens | 130,856 | 22,400 | 413,231 |

| Characteristic | AM dataset | EVD-100 | STEMI |
| --- | --- | --- | --- |
| Number of records | 405 | 100 | 1730 |
| Avg. words per anamnesis | 256 (12–1,113) | 188 (7–383) | 202 (2–348) |
| Avg. characters per anamnesis | 1,869 (70–8,206) | 1,357 (47–2,517) | 1,455 (29–2,047) |
| Total sentences | 11,469 | 1,520 | 28,956 |
| Total tokens | 130,856 | 22,400 | 413,231 |

### 3.2 Annotation and tokenization for NER

We used Label Studio [21] to manually annotate anamneses with clinical tags crucial for assessment (Fig. 1b). These include hypertension (HY/NOHY), dyslipidemia (DY/NODY), diabetes (DB/NODB), COPD (COPD/NOCOPD), NYHA class (1–4), ejection fraction (EF, 0–100), sinus rhythm (RS/NORS), atrial fibrillation/flutter (FA/NOFA, FL/NOFL), pacemaker presence (PM/NOPM), and bundle branch blocks (BBSx/NOBBSx, BBDx/NOBBDx). All features are boolean except NYHA and EF, which are integers.

The annotations were converted to Inside-Outside-Beginning (IOB) format [22] and tokenized using an Italian model [23] pre-trained on ISDT, VIT, POSTWITA, and TWITTIRO within Stanza [24]. Each token was labeled with a clinical tag or marked ‘O’ if irrelevant.

For each clinical feature, we implemented a three-class classification scheme:

1. **Presence** – the variable is explicitly documented (e.g., “diabete mellito”)
2. **Absence** – the variable is explicitly negated (e.g., “no diabetes”, “nega diabete”)
3. **Not mentioned** – the variable does not appear in the EHR narrative

This distinction is critical: features not mentioned reflect clinicians’ selective documentation practices where features are recorded only when clinically relevant, not missing data. Performance metrics are calculated exclusively on mentioned features (Presence or Absence), as ground truth annotations only exist for documented features.

### 3.3 NER for anamnesis structuring

As presented in the following sections, we investigated one string-based baseline approach (Section 3.3.1) and three NER models: Multiconer (Section 3.3.2), SpaCy (Section 3.3.3) and Flair (Section 3.3.4).

#### 3.3.1 Baseline

We implemented a string-based NER baseline without transformers, consisting of dictionary construction and prediction. This baseline identifies clinical features explicitly mentioned in the text (Presence or Absence). Dictionaries map entity tags to lower-cased forms from input sequences. During prediction, subsequences in sentences are matched to tags via a reverse dictionary, applying non-overlapping tags sequentially. Input data are in IOB format. While efficient, this method lacks ability to handle language variability and context, limiting accuracy compared to transformer-based models. Detailed information about multi-token dictionary construction, matching procedures, and lexical variants is provided in the Appendix (Table 2).

#### 3.3.2 Multiconer

Multiconer [25] uses token embeddings to capture context and applies a Conditional Random Field (CRF) for sequence labeling. Built on a pretrained encoder (e.g., XLM-RoBERTa-Large [26]), it includes a feedforward layer for token classification. Training employed AdamW optimizer with a learning rate of 0.0001, early stopping after 10 epochs without improvement, batch size 64, and negative log likelihood loss.

#### 3.3.3 SpaCy

We customized SpaCy2 [27] by integrating a BERT-based language model pretrained on OPUS and OSAMR Italian corpora. SpaCy’s tokenizer and transition-based parser handle tokenization and NER, respectively. Training used multi-label log loss, Adam optimizer with warmup linear scheduling, patience 5600 steps, batch size 128, gradient accumulation over 3 steps, and L2 regularization to reduce overfitting.

#### 3.3.4 Flair

Flair [28] employs the ‘dbmdz/bert-base-italian-xxl-cased’ BERT model to generate contextual embeddings, combined with a CRF layer using Viterbi loss for sequence labeling. Training was run for 200 epochs with a learning rate of 5e-5, batch size 64, and warmup linear scheduling. Dropout techniques regularized the model and improved prediction accuracy.

### 3.4 Experimental protocol

The AM dataset was split with 80% for training (further split 80/20 into train/validation) and 20% for testing, totaling 6790 training, 1886 validation, and 2239 test sentences.

NER hyperparameters were chosen experimentally without further tuning. Performance was evaluated using True Positives (TP), True Negatives (TN), False Positives (FP), False Negatives (FN), and metrics Precision (P_nlp), Recall (R_nlp), and F1-score (F1_nlp). TP means correct clinical feature detection; TN means correct identification of absence of that features; FP and FN indicate incorrect tagging or missed entities, respectively.

External validation used STEMI and EVD-100 datasets, comparing only entities common with AM that for STEMI analysis focused on hypertension, dyslipidemia, and diabetes.

All performance metrics (Precision, Recall, F1-score) are calculated exclusively on instances where clinical features are mentioned in the text (Presence or Absence cases). This methodological choice reflects the fact that ground truth annotations are only available for documented features. Features not mentioned in the clinical narrative are not included in performance calculations, as there is no clinician annotation to compare against. This approach accurately reflects the practical utility of the NER model: its ability to correctly identify and classify features when they are documented by clinicians.

For downstream classification of AM in heart failure patients, structured anamneses from the NER test set (83 cases) were used. We employed Logistic Regression (LR), XGBoost (XGB), and CatBoost (CAT), chosen for effectiveness in clinical settings and small datasets. We prioritized conventional models due to their competitiveness in disease onset prediction, especially with small datasets, despite the rising capabilities of deep learning in structured healthcare data [29]. As a baseline, we used LR for its interpretability and effectiveness in modeling approximately linear relationships [30]. We also employed XGB, a boosting algorithm known for reducing false positives [31], and CAT, which handles categorical features efficiently and performs well across various clinical scenarios [32].

The downstream classification task serves dual purposes: (1) showing that automatically extracted clinical features contain sufficient predictive information for machine learning applications, and (2) validating whether NER-extracted features can substitute manual annotation without compromising classifier performance. This comparison addresses a critical question for clinical NLP adoption: can automated systems replace time-intensive manual annotation while maintaining equivalent predictive accuracy?

A Monte Carlo cross-validation with 10 iterations and 80/20 train/test splits (66/17 samples) was used. Models were compared via Accuracy (A_cl), Precision (P_cl), Recall (R_cl), and F1-score (F1_cl), where TP, TN, FP, and FN denote the correct and incorrect predictions regarding the presence of AM.

Hyperparameters were tuned via grid search: LR with L2 penalty and *C* ∈ {0.1, 1}; XGB with max depth {3, 5}, estimators {50, 100}, learning rates {0.01, 0.1}, feature/sample fractions; CAT with iterations {100, 200}, depth {4, 6}, learning rates {0.01, 0.05}, and regularization {1, 5}. Feature importance was analyzed via Gini index; precision-recall curves were examined.

Normality and homogeneity of variance were verified with Shapiro Wilk and Levene’s tests. Repeated measures ANOVA assessed significant differences in classifier performance across datasets (NER-AM, GoldStd) and models (XGB, CAT, LR).

The code is publicly available at: https://github.com/saramazz/NLP_Italian_EHRs.git

## 4 Results

From 405 EHRs in the AM dataset, we extracted 10,906 sentences and 25,920 tokens. Figure 3 (a) displays the distribution of named clinical features detected by NLP models.

### 4.1 NER performance

Figure 4 reports test set performance of the baseline and three NER models, showing precision, recall, and F1-score for each clinical feature (including negations) and overall metrics. The Appendix includes confusion matrices (Fig. 8) and validation set results (Table 3) for the NER models used.

#### 4.1.1 Performance on Mentioned Entities

The overall F1-score of 97% represents performance on the subset of clinical features that are explicitly mentioned in the EHR narratives. As shown in Figure 3(a), there are a lot of “not mentioned” values across the dataset. This reflects real-world clinical documentation patterns where features are selectively recorded based on clinical relevance (e.g., ejection fraction typically documented when abnormal or routinely measured, NYHA classification documented when dyspnea is present). On the evaluated subset of mentioned features, the model achieves consistent high performance across all clinical categories.

#### 4.1.2 Validation on external datasets

External validation was conducted on two datasets with different annotation coverage, reflecting real-world availability of annotated resources.

The EVD-100 dataset provided clinician annotations for all 12 clinical features, enabling comprehensive validation of the complete model. SpaCy achieved 97.13% F1-score with 97.13% alignment with clinician labels across all features. Figure 5 shows the F1-scores per label across datasets.

**Fig. 3.**
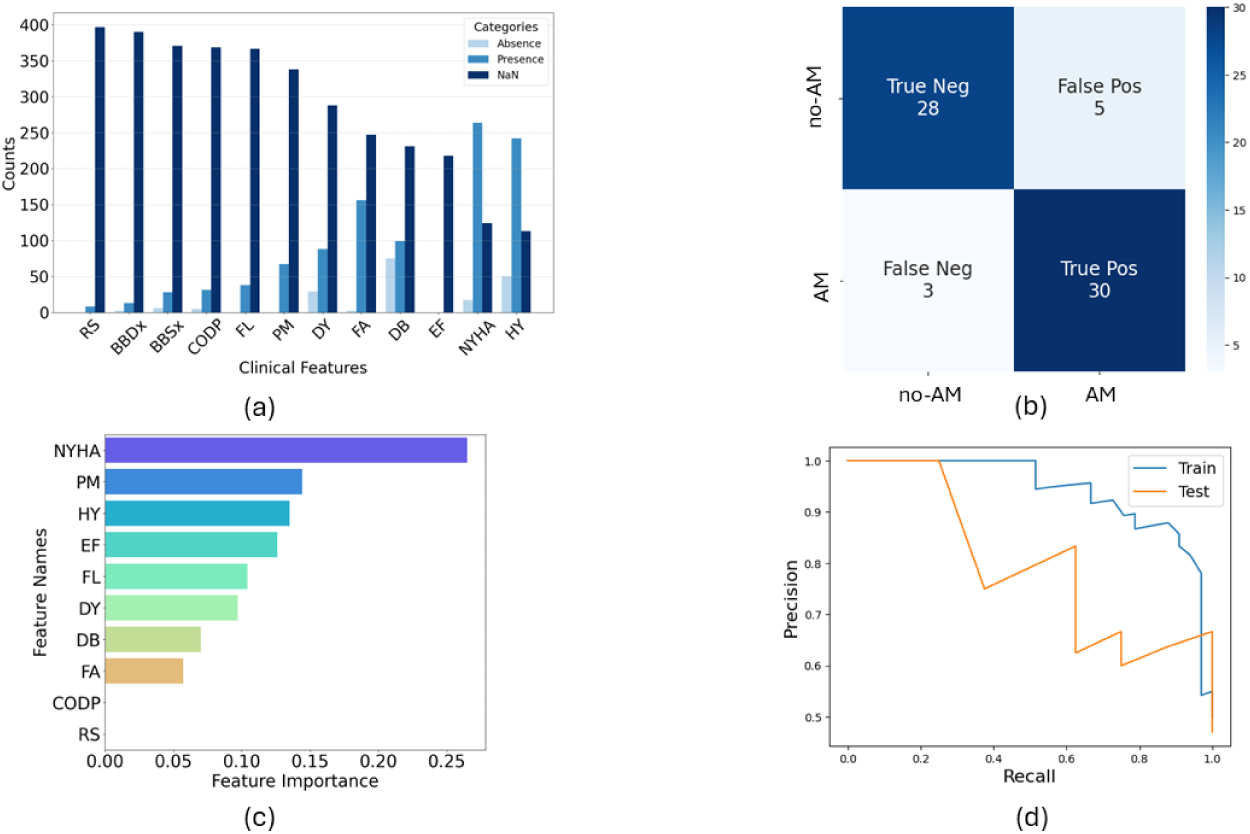
(a) Distribution of values for each clinical feature extracted from the NER model in the AM dataset. The plot categorizes data into three classes: Presence (features explicitly documented), Absence (explicit negation of features), and NaN (not mentioned in the clinical text). Note that “NaN” does not represent corrupted or incomplete data, but rather clinical information not recorded by clinicians, reflecting selective documentation patterns in real clinical practice. (b) Confusion Matrix, (c) Top 10 Feature importance. (d) Precision-Recall curves for AM detection from NER-AM dataset for XGBoost (XGB) classifier. All the acronyms are detailed in Fig. 1b

Among 29 mismatches, the distribution was DY 14%, HY 6%, DB 3%, CODP 3%, NYHA 1%, BBSx 1%, BBDx 1%. 76% of mismatches were due to clinicians inferring features from medication rather than explicit EHR mentions, indicating high agreement between NER and clinician judgment.

We also validated on the STEMI dataset. However, this dataset was annotated by external centers for only 3 features (hypertension, dyslipidemia, diabetes). SpaCy achieved 88.29% F1-score on these shared features. The lower performance compared to EVD-100 may reflect differences in clinical populations and documentation patterns in STEMI versus amyloidosis settings.

### 4.2 Downstream task performance

Figure 6 shows accuracy, precision, recall, and F1-score of classifiers on the NER-AM dataset (SpaCy-annotated) and GoldStd dataset (clinician-annotated). XGB was best on NER-AM with 71.43% F1, close to 70.59% on GoldStd. Two-way ANOVA found no significant difference in F1 between datasets or classifiers (*p >* 0.05), suggesting that the NER pipeline achieves annotation performance equivalent to that of clinical experts in classifying amyloidosis in heart failure patients.

**Fig. 4.**
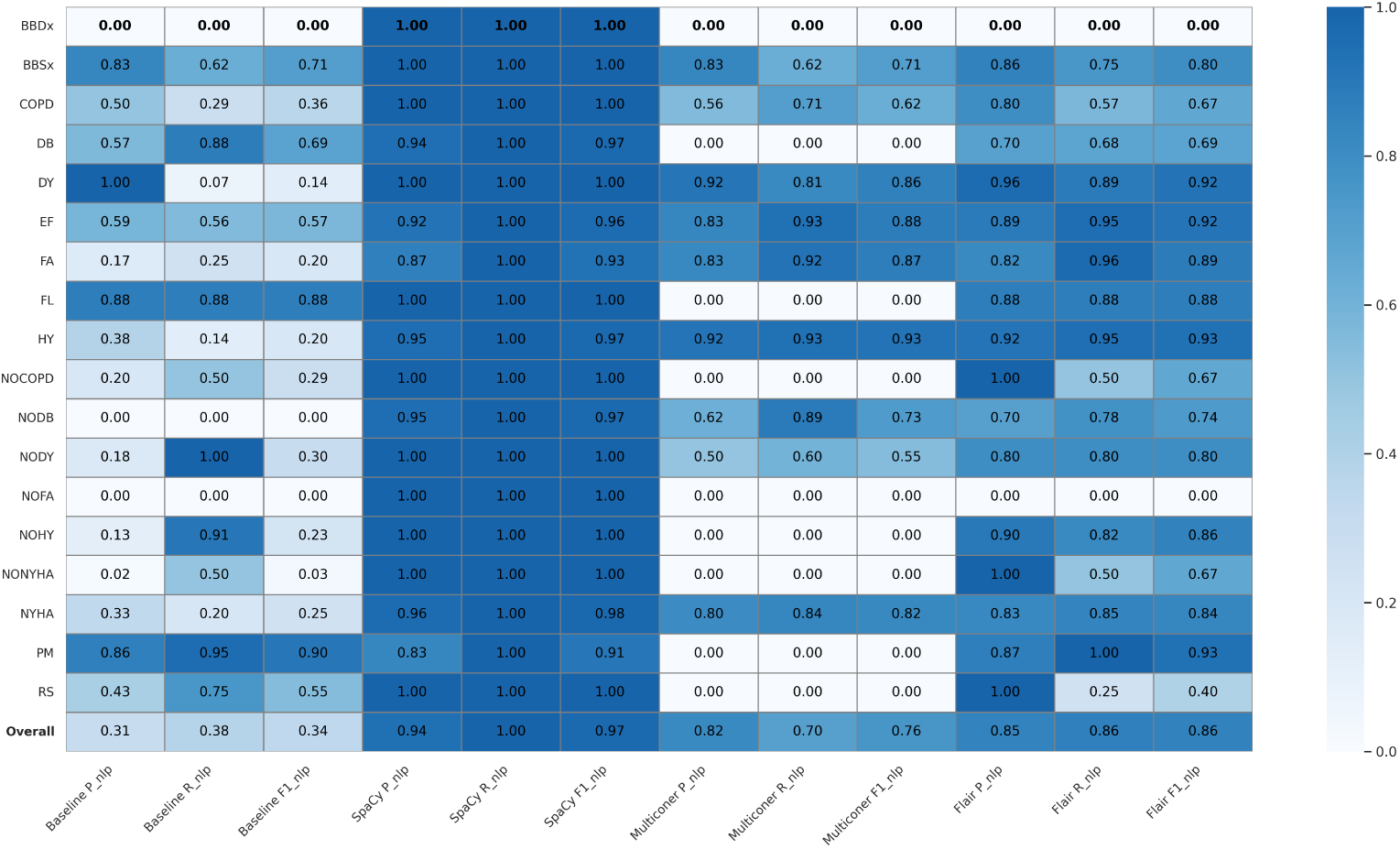
NER Results on the AM dataset: Precision (P_nlp), Recall (R_nlp), and F1-score (F1_nlp) of Baseline vs. SpaCy vs. Multiconer vs. Flair

**Fig. 5.**
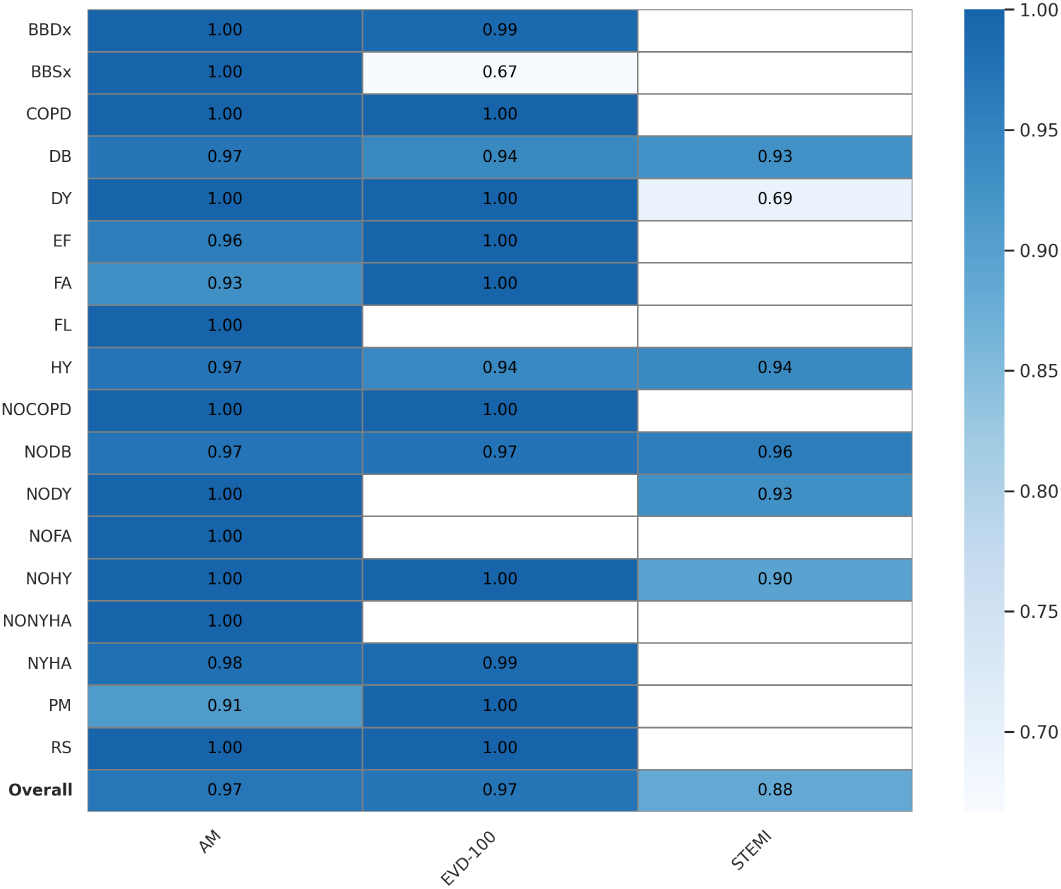
F1-score for the recognition of clinical features across the three datasets. Scores are reported only for labels that were manually annotated in each dataset; cells are left blank where annotations were not available

**Fig. 6.**
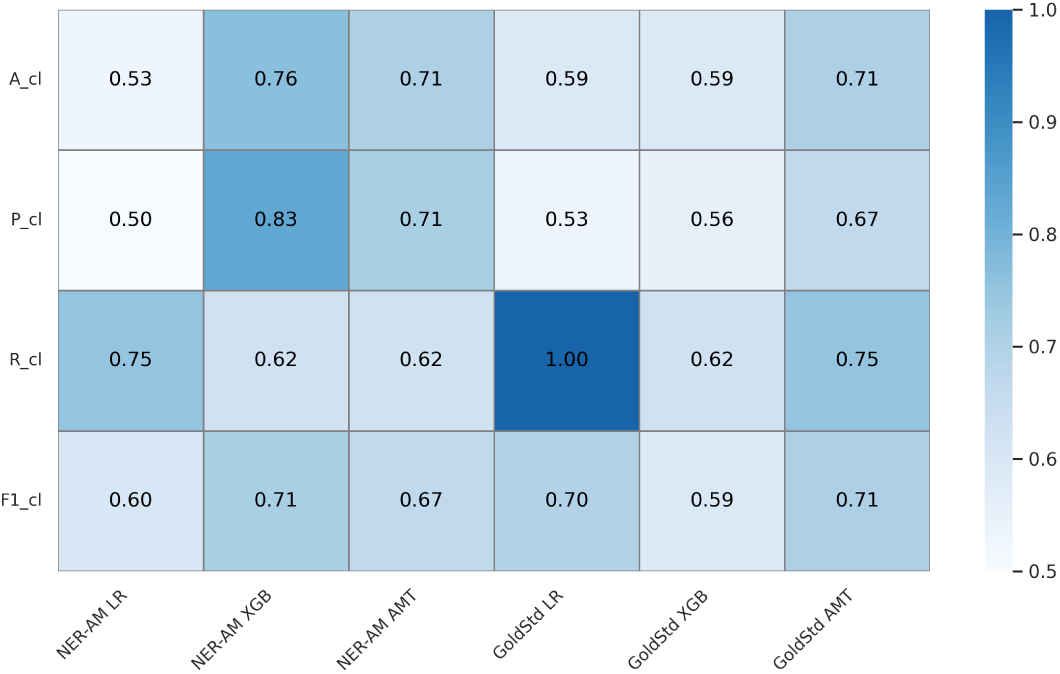
Accuracy (A_cl), Precision (P_cl), Recall (R_cl), and F1-score (F1_cl) obtained with the three classifiers for detection of amyloidosis using the NER-AM and GoldStd datasets on the test set

Panels (b), (c), and (d) of Fig. 3 display the classification confusion matrix, top 10 feature importance, and precision-recall curves for AM detection using XGB on NER-AM, offering a detailed model evaluation.

## 5 Discussion

In this study, we introduced a systematic framework that utilized anamneses from EHRs, integrating free-text information. Despite its value, unstructured data sources like free-text often remain underutilized. We evaluated our NER pipeline across three real-world EHR databases for automatically identifying clinical features of interest. Our study contributes by evaluating NLP approaches for Italian data, representing the first attempt to develop a pipeline for Italian cardiology EHRs extracting a comprehensive set of clinical features, with applicability across multiple domains, as shown on the EVD-100 multidomain dataset. This addresses challenges in NLP for EHR structuring, particularly pipeline integration and scarcity of non-English datasets.

The NER model showed promising results, with SpaCy as the best approach, achieving an F1-score of 96.93% on the AM dataset. This is due to SpaCy’s pretrained models on large datasets, enhancing baseline NLP performance. Its architecture is designed for speed and efficiency, supporting large datasets. SpaCy’s multilingual and medical language processing models expanded the scope of our work, making it more versatile than FLAIR and Multiconer XML-RoBERTa. The flexibility allowed accurate identification of clinical features in Italian EHRs. Performance varied by feature frequency; features like FA (72% not mentioned) and PM (87% not mentioned) had F1 around 91%. Optimizing parameters might improve results, especially for Multiconer. Compared to state of the art models like GCT and LSTMs [11, 13], our approach leverages transformers’ strengths in handling complex texts. Unlike GCT, which may falter on concise texts, and LSTMs, which struggle with sporadic features, our method robustly structures variable-length cardiac anamneses. While Garvin et al.[15] achieved high accuracy for a single feature (EF), our approach handles multiple features.

On the external STEMI dataset, the model scored 88.29% F1, showing in 4 good agreement with manual annotations. The lower score versus AM data may stem from clinical profile differences and more not mentioned values in STEMI records, 66.82% for DB, 66.30% for DY, and 36.59% for HY, since these features are less common in STEMI histories. The pipeline extracted clinical labels like NYHA, previously unstructured in the registry, enabling future research on disease progression. NYHA had fewer not mentioned values (63.12%) compared to DY and DB, which are often left unfilled by clinicians.

The EVD-100 dataset yielded strong performance (F1 = 97.13%), with mismatches mostly due to physician inference from drug therapy, beyond model scope.

An important clarification regarding the high proportion of “not-mentioned” features (Figure 3a): these do not represent data quality issues or incomplete records, but rather reflect clinician documentation practices in real-world clinical settings. Clinicians selectively record features based on clinical relevance and workflow constraints. This selective documentation is a feature of real clinical practice, not a limitation of our dataset. Consequently, our performance metrics appropriately evaluate the model only on documented features, where ground truth annotations exist, achieving 97% F1-score on these evaluated instances.

Regarding patient status classification in the GoldStd and NER-AM datasets, XGB achieved F1-scores of 71.43% and 70.59%, respectively, with no statistically significant difference (*p >* 0.05). This comparable performance shows that our NLP pipeline can effectively substitute time-intensive manual annotation without compromising predictive accuracy. The finding has important practical implications: automated feature extraction enables scalable, real-time clinical decision support systems for risk stratification and patient monitoring, while substantially reducing the cost and effort of manual EHR annotation.

Feature importance varied between classifiers, reflecting algorithm-specific strategies and the fact that not all features are causative of AM. In XGB, key variables included NYHA, PM, HY, EF, FA, DY, and DB. These align with literature: NYHA reflects patient functionality [33]; PM relates to advanced dysfunction [34]; HY indicates early cardiovascular issues [35]; EF distinguishes amyloid cardiomyopathy [36]; FA increases arrhythmic risk [37].

Conversely, features like CODP and RS had limited impact, likely due to weak cardiac specificity and high not mentioned rates. While such variables support generalization, heart-specific markers remain essential for precise diagnosis and management in amyloidosis.

### 5.1 Limitations and future work

This study is limited by the scarcity and limited diversity of annotated data, though its size is comparable to similar research [38]. Although we employed general cardiac risk factors rather than amyloidosis-specific features due to limited available data, this approach provides broader generalizability across cardiac conditions.

The extraction of 12 clinical features, predominantly boolean variables, represents a focused but limited subset of potentially relevant clinical information. However, these features were selected based on established clinical literature for cardiac risk stratification [20], ensuring clinical relevance. Future work will expand the feature space to include continuous variables and disease-specific markers such as carpal tunnel syndrome and spinal stenosis.

The high proportion of not-mentioned features (Figure 3a) reflects selective clinical documentation practices rather than data quality issues, as physicians record features based on clinical relevance. While this limits assessment of complete patient profiles, our performance metrics appropriately evaluate the NER model only on documented features, achieving 97% F1-score. Moreover, external validation mitigates this concern: EVD-100 validates all 12 features comprehensively (97.13% F1), while STEMI validates 3 shared features (88.29% F1) despite annotation constraints imposed by external data availability rather than model limitations.

Our approach analyzes single-timepoint clinical summaries rather than longitudinal records. While the NER model effectively extracts documented features, it does not capture temporal disease trajectories or comorbidity development over time. Future work could extend this to longitudinal EHR data from sequential encounters, enabling analysis of temporal patterns in disease progression through appropriate temporal modeling techniques.

Future work includes employing generative models to augment training data [39], particularly for rare clinical expressions. We also plan to integrate explainability techniques to improve model interpretability, conduct multi-center validation studies with complete feature annotations, and compare our approach with open-source large language models. Further studies will explore applying the model to other pathologies. Despite these limitations, our work demonstrates the value of NER methods in automatically extracting clinical information from Italian EHRs, achieving performance comparable to manual annotation while significantly reducing manual workload in healthcare.

## 6 Conclusion

This study investigates the potential of NLP in structuring Italian EHRs for cardiac settings, particularly in AM detection, exploring three transformer-based NER algorithms, SpaCy, Flair, and Multiconer, with SpaCy achieving a recall of 100% on the AM dataset. Using Italian datasets from FTGM, we employed SpaCy’s structured data for identifying cardiac amyloidosis and heart failure through machine learning algorithms. XGBoost emerged as the most effective classifier, achieving a 71% F1-score on the cardiac amyloidosis dataset. This shows the robustness of our NLP pipeline and its comparable effectiveness to traditional clinician-annotated datasets. These findings support the integration of automated NLP methods into clinical workflows, potentially streamlining processes and conserving resources. Implementing our pipeline in clinical settings for real-time validation could revolutionize practice, offering prompt and accurate insights into patient conditions. This research addresses challenges in managing unstructured digital records, especially in non-English languages, enhancing healthcare efficiency and paving the way for future advancements in using NLP to improve patient care and disease detection.

## Data Availability

The data that support the findings of this study are not publicly available due to ethical and legal restrictions, but are available from the corresponding author upon reasonable request and with approval from the Ethics Committee.

## Declarations

### Ethics approval and consent to participate

The study was approved by the FTGM Ethics Committee (Decree No. 3854, 02/12/2023) and was conducted in accordance with the Declaration of Helsinki. All participants provided informed consent. Clinical records were anonymized at source by FTGM staff prior to data sharing, following GDPR-compliant protocols.

### Consent for publication

Written informed consent for publication was obtained from all participants. No identifiable patient data, images, or case details are included in this manuscript. All procedures were conducted in accordance with COPE guidelines.

### Availability of data and material

The data underlying this study cannot be shared publicly due to patient privacy and ethical restrictions. The code for data processing and analysis is available at https://github.com/saramazz/NLP_Italian_EHRs.git.

### Competing interests

The authors declare that they have no competing interests.

## Funding

No funding was received.

## Authors’ contributions

S. Mazzucato performed the data analysis, drafted the original manuscript, and implemented all subsequent revisions. A. Bandini acted as supervisor and supervised the manuscript draft. D. Sartiano contributed to data analysis and manuscript revision. G. Vergaro provided the clinical data. S. Dalmiani provided the clinical data. M. Emdin coordinated the project and contributed to funding acquisition. S. Micera coordinated the project and contributed to funding acquisition. C. M. Oddo contributed to supervision and manuscript review. C. Passino coordinated the project, contributed to funding acquisition, and reviewed the manuscript. S. Moccia contributed to supervision and manuscript review. All authors read and approved the final manuscript.

## Acknowledgements

We thank the “Proximity Care Project” by Scuola Superiore Sant’Anna – Interdisciplinary Center “Health Science,” supported by Fondazione Cassa di Risparmio di Lucca, and collaborators including Azienda USL Toscana Nord Ovest and Fondazione Monasterio.

## Clinical Trial Number

Not applicable.

## Appendix

### 6.1 Dataset Description: Distribution of anamnesis lengths

An overview of the distribution of anamnesis lengths across the three datasets is provided in Figure 7.

### 6.2 Baseline Dictionary for Multi-Token Matching

The baseline employs a multi-token sequence matching approach using hand-crafted dictionaries extracted from annotated clinical data. The complete dictionary (baseline dictionary v2.json) contains 12 clinical condition categories plus negation classes (288+ variants total) and is provided here and in the git repository for reproducibility.

#### 6.2.1 Matching Algorithm and Implementation

The dictionary uses **longest-match-first** strategy with exact string matching (case-insensitive). Preprocessing includes lowercase conversion, whitespace normalization, and BIO tagging (B- for entity start, I- for continuation). Sequences are processed in descending length order to ensure longer (more specific) matches are preferred, with a non-overlapping constraint preventing span reuse.

**Fig. 7.**
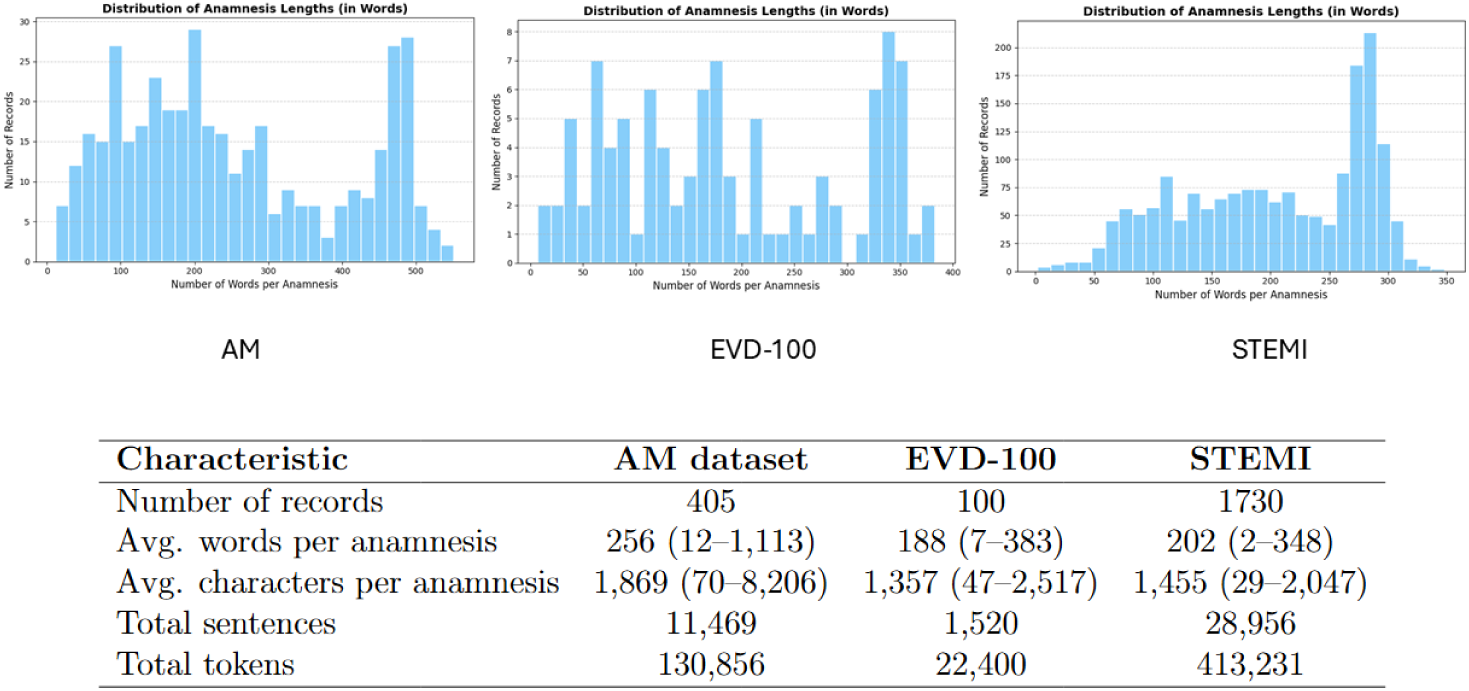
Distribution of anamnesis lengths (in words) across the three datasets. (a) CA dataset: average 256 words (range: 12–1,113), 11,469 sentences, 130,856 tokens. (b) EVD-100 dataset: average 188 words (range: 7–383), 1,520 sentences, 22,400 tokens. (c) STEMI dataset: average 202 words (range: 2–348), 28,956 sentences, 413,231 tokens. The distributions demonstrate substantial variability in clinical documentation practices across different cardiac conditions and clinical settings.

Dictionary construction involved extracting multi-token sequences from annotated training data, grouping by clinical label, and retaining all unique surface forms including spelling variants, abbreviations, and misspellings. Negation categories (B-NO* prefix) explicitly mark absence of clinical conditions.

For reproducibility:

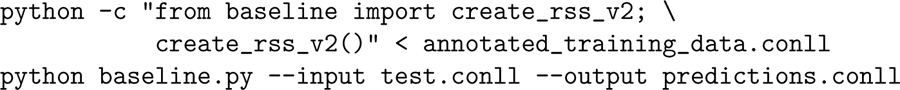

#### 6.2.2 Dictionary Statistics and Format

The composition of the multi-token clinical dictionary, including the number of variants for each clinical category and the associated negation classes, is summarized in Table 2.

#### 6.2.3 Complete Dictionary

The full dictionary in JSON format:

**Table 2.** Multi-token dictionary: 12 clinical variables plus negation classes.

| Category | Clinical Condition | Variants |
| --- | --- | --- |
| B-HY | Hypertension | 13 |
| B-DB | Type 2 Diabetes | 6 |
| B-FA | Atrial Fibrillation | 9 |
| B-EF | Ejection Fraction | 76 |
| B-FL | Atrial Flutter | 2 |
| B-RS | QRS Complex | 3 |
| B-DY | Dyslipidemia | 5 |
| B-NYHA | NYHA Classification | 128 |
| B-BBSx | Left Bundle Branch Block | 6 |
| B-BBDx | Right Bundle Branch Block | 3 |
| B-PM | Pacemaker | 5 |
| B-CODP | COPD | 5 |
| B-NO* | Negation classes | 38 |
| **Total** |  | **299+** |

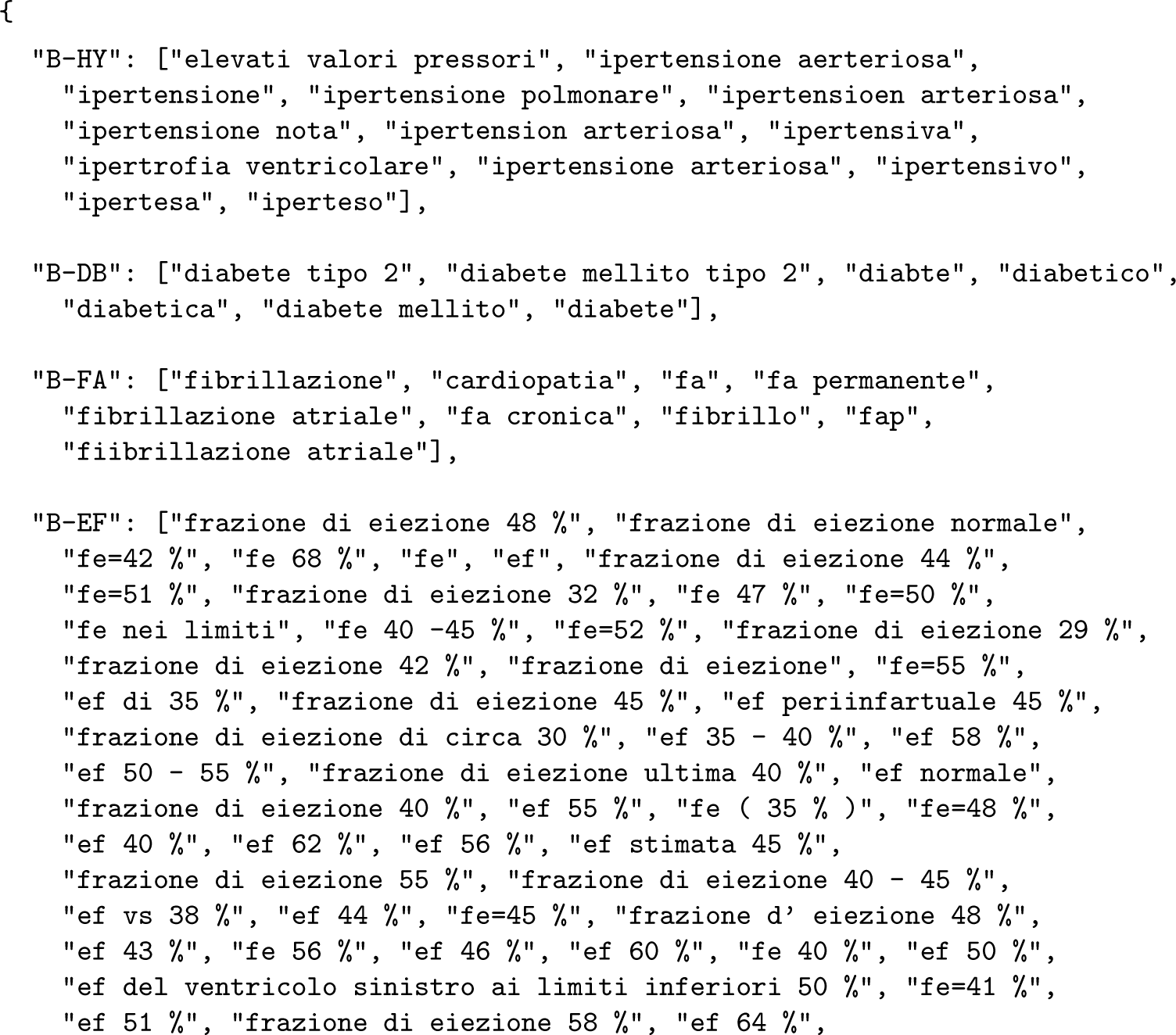
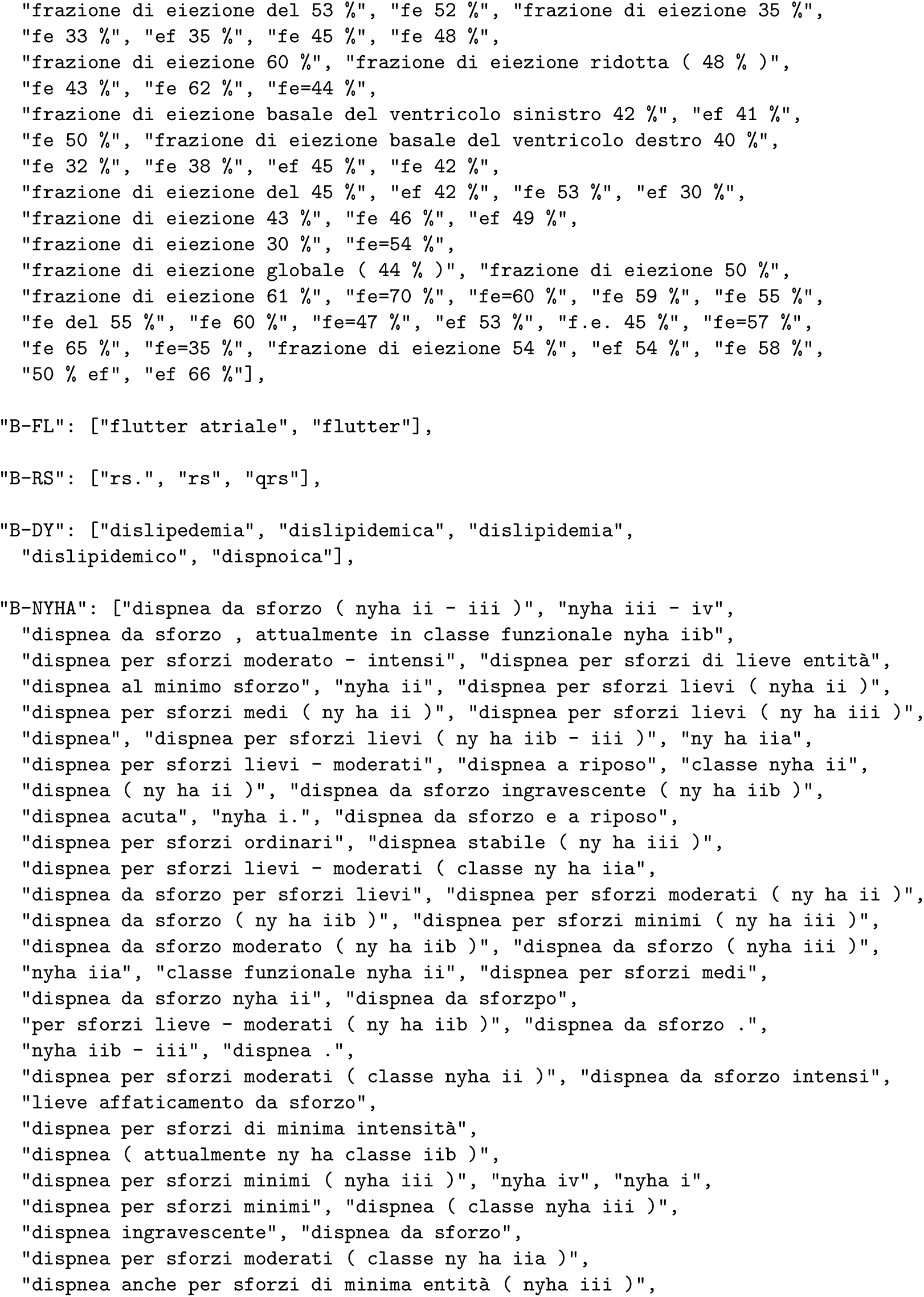
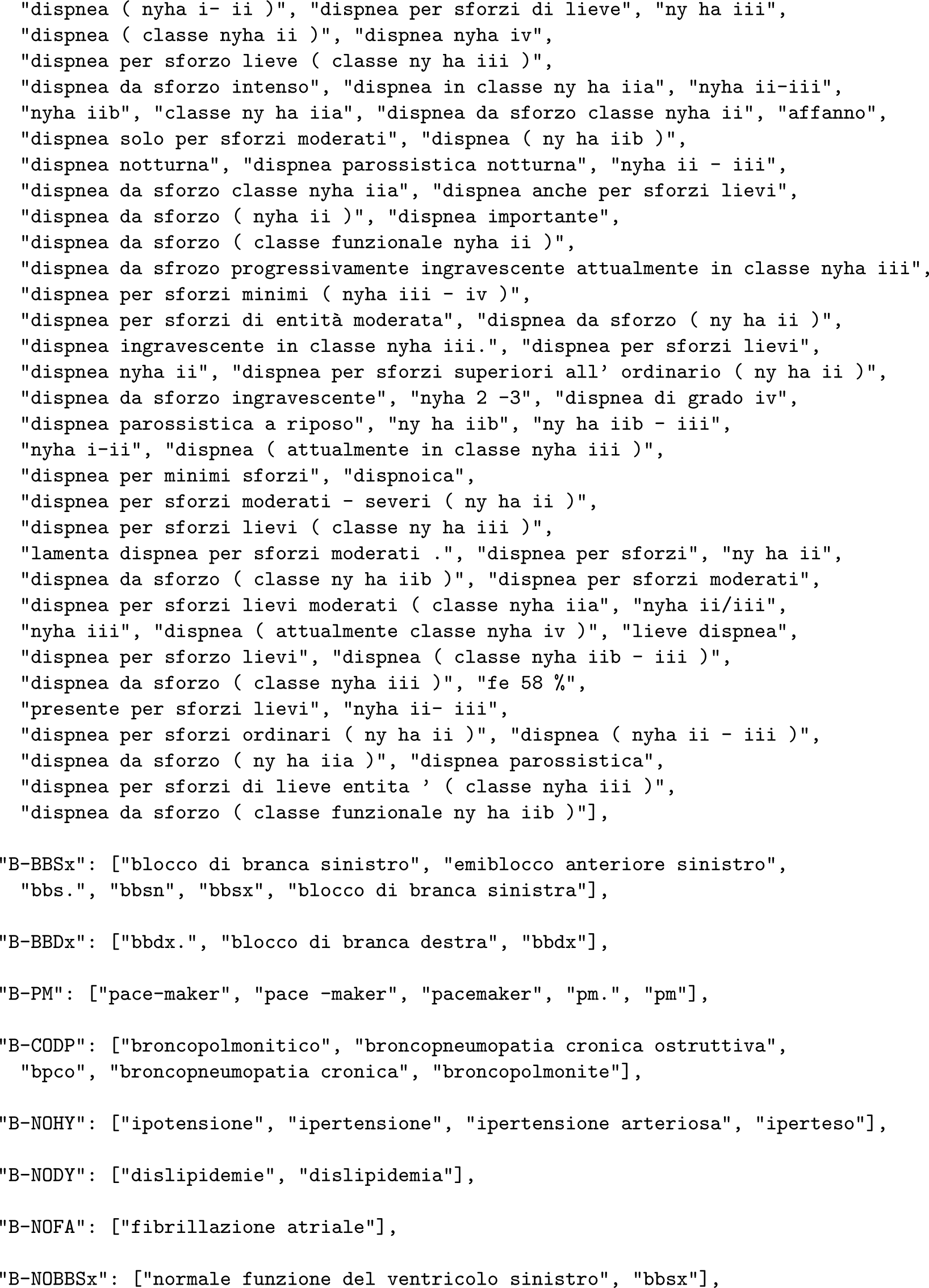
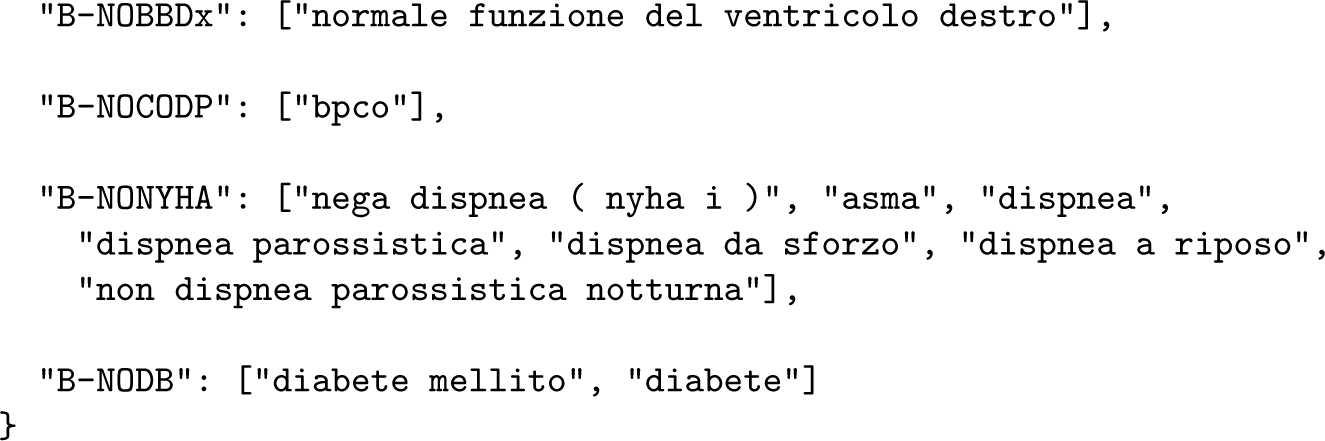

### 6.3 NER model validation performance

Table 3 shows NER model validation performance used for model selection.

**Table 3.** NER Results on the AM dataset: Precision (P_nlp), Recall (R_nlp), and F1-score (F1_nlp) of Baseline vs. SpaCy vs. Multiconer vs. Flair on the validation Set.

|  | Baseline |  |  | SpaCy |  |  | Multiconer |  |  | Flair |  |  |
| --- | --- | --- | --- | --- | --- | --- | --- | --- | --- | --- | --- | --- |
|  | P<sub>nlp</sub> | R<sub>nlp</sub> | F1<sub>nlp</sub> | P<sub>nlp</sub> | R<sub>nlp</sub> | F1<sub>nlp</sub> | P<sub>nlp</sub> | R<sub>nlp</sub> | F1<sub>nlp</sub> | P<sub>nlp</sub> | R<sub>nlp</sub> | F1<sub>nlp</sub> |
| BBDx | 100.00% | 75.00% | 85.71% | 100.00% | 100.00% | 100.00% | 100.00% | 50.00% | 66.67% | 100.00% | 50.00% | 66.67% |
| BBSx | 75.00% | 100.00% | 85.71% | 100.00% | 100.00% | 100.00% | 25.00% | 33.33% | 28.57% | 25.00% | 33.33% | 28.57% |
| COPD | 0.00% | 0.00% | 0.00% | 85.71% | 100.00% | 92.31% | 50.00% | 66.67% | 57.14% | 71.43% | 83.33% | 76.92% |
| DB | 62.79% | 87.10% | 72.97% | 93.94% | 100.00% | 96.88% | 0.00% | 0.00% | 0.00% | 74.19% | 74.19% | 74.19% |
| DY | 100.00% | 9.09% | 16.67% | 100.00% | 100.00% | 100.00% | 95.45% | 95.45% | 95.45% | 100.00% | 100.00% | 100.00% |
| EF | 63.24% | 65.15% | 64.18% | 94.29% | 100.00% | 97.06% | 91.30% | 95.45% | 93.33% | 87.14% | 92.42% | 89.71% |
| FA | 8.33% | 10.00% | 9.09% | 97.56% | 100.00% | 98.77% | 92.68% | 95.00% | 93.83% | 90.91% | 100.00% | 95.24% |
| FL | 100.00% | 100.00% | 100.00% | 100.00% | 100.00% | 100.00% | 0.00% | 0.00% | 0.00% | 85.71% | 100.00% | 92.31% |
| HY | 52.17% | 17.65% | 26.37% | 97.14% | 100.00% | 98.55% | 96.77% | 88.24% | 92.31% | 95.31% | 89.71% | 92.42% |
| NOCOPD | 14.29% | 50.00% | 22.22% | 100.00% | 100.00% | 100.00% | 0.00% | 0.00% | 0.00% | 100.00% | 50.00% | 66.67% |
| NODB | 0.00% | 0.00% | 0.00% | 100.00% | 100.00% | 100.00% | 52.94% | 90.00% | 66.67% | 80.00% | 80.00% | 80.00% |
| NODY | 16.67% | 100.00% | 28.57% | 100.00% | 100.00% | 100.00% | 75.00% | 75.00% | 75.00% | 100.00% | 100.00% | 100.00% |
| NOFA | 0.00% | 0.00% | 0.00% | 0.00% | 0.00% | 0.00% | 0.00% | 0.00% | 0.00% | 0.00% | 0.00% | 0.00% |
| NOHY | 20.83% | 88.24% | 33.71% | 85.00% | 100.00% | 91.89% | 0.00% | 0.00% | 0.00% | 82.35% | 82.35% | 82.35% |
| NONYHA | 4.44% | 66.67% | 8.33% | 100.00% | 100.00% | 100.00% | 0.00% | 0.00% | 0.00% | 28.57% | 66.67% | 40.00% |
| NYHA | 44.83% | 30.59% | 36.36% | 94.44% | 100.00% | 97.14% | 85.19% | 81.18% | 83.13% | 85.19% | 81.18% | 83.13% |
| PM | 80.00% | 100.00% | 88.89% | 70.59% | 100.00% | 82.76% | 0.00% | 0.00% | 0.00% | 85.71% | 100.00% | 92.31% |
| RS | 0.00% | 0.00% | 0.00% | 0.00% | 0.00% | 0.00% | 0.00% | 0.00% | 0.00% | 0.00% | 0.00% | 0.00% |
| Overall | 34.86% | 41.78% | 38.00% | 93.87% | 100.00% | 96.84% | 87.10% | 70.50% | 77.92% | 85.31% | 86.42% | 85.86% |

### 6.4 NER confusion matrices

Figure 8 presents confusion matrices for entity recognition.

**Fig. 8.**
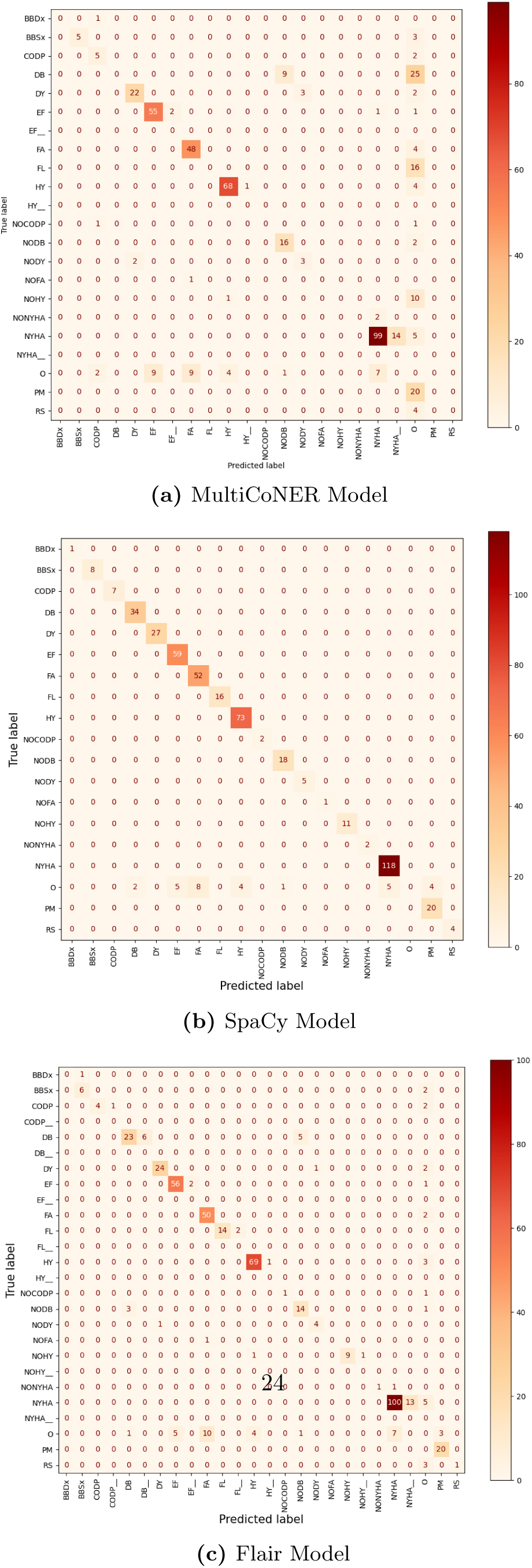
Confusion Matrices for Different NER Models. Each subfigure displays the performance of a different model. Labels with a ‘–’ suffix indicate partial span and structure agreement

## Footnotes

1 https://labelstud.io/

## Notes

### Competing Interest Statement

The authors have declared no competing interest.

### Funding Statement

This study did not receive any funding

### Author Declarations

Ethics Committee of Fondazione Toscana Gabriele Monasterio gave ethical approval for this work (Decree No. 3854, 02 December 2023).

